# Social prescribing for children and young people in the UK: characterising access and care pathways using electronic health records

**DOI:** 10.64898/2026.06.02.26354692

**Authors:** Jessica K Bone, Feifei Bu, Daniel Hayes, Daisy Fancourt

## Abstract

**Objectives:** We aimed to describe the characteristics of children and young people referred to social prescribing across the UK and understand what social prescribing looks like for these young people. Additionally, we aimed to explore whether access to and experiences of social prescribing vary with age and have changed from 2017 to 2025. Overall, we aimed to identify whether social prescribing reduces or exacerbates health inequalities among children and young people, and whether this has changed over time.

**Design:** Analysis of social prescribing electronic health records

**Setting:** Social prescribing hubs and services across the UK that use Access Elemental (a cloud-based social prescribing platform)

**Participants:** 52,585 individuals referred to social prescribing in 2017-2025 aged 4-25 years (mean=20.04, SD=4.71), of whom 57% were female, 39% male, <2% were in other gender groups, and 3% did not disclose their gender

**Primary and secondary outcome measures:** We summarised young people’s characteristics and described the care pathway received. We then used regression models to test whether these factors differed by age and over time.

**Results:** Most individuals were aged 18 and over, 91% lived in urban areas and 58% lived in the top three most deprived deciles of the UK. Most were referred by GPs or other allied health workers (79%) and mental health was the leading reason for referral (44%). The typical pathway included 4.64 social prescribing contacts (SD=7.70) totalling 66 minutes (SD=108), with 34% receiving an onward referral to community support. The average age of those referred to social prescribing increased over time.

**Conclusions:** Our findings indicate that social prescribing currently has limited reach for those under 18 and this disparity may be increasing. It was promising that children and young people referred to social prescribing were more likely to live in deprived areas. However, given current findings, more work is needed to increase the reach of social prescribing for children and young people across the UK.

**Strengths and limitations of this study:**

- We used data from the most widely adopted social prescribing platform in the UK, including a large and diverse population of children and young people
- We included young people referred to social prescribing through medical and non-medical pathways, which is key for understanding inequalities in service provision
- Administrative data were limited by missingness and varying data quality, meaning we could not explore the role of ethnicity among other factors
- It was unclear whether individuals with no recorded interventions did not receive any prescriptions or were missing these data
- We could only explore access to and delivery of social prescribing by sites that have chosen to use the Access Elemental platform, making it difficult to understand what was driving changes over time

## Introduction

In the UK, there is rising demand for health services, with an increasing number of children and young people (CYP) experiencing poor physical and mental health.^1–3^ Current services do not have capacity to serve these CYP. The 10 Year Health Plan for England aims to address these challenges through community-based approaches to prevention, shifting care into the community and addressing the wider determinants of health.^4^ Social prescribing (SP), a care pathway that aims to connect people with non-medical forms of support within their community based on their values and preferences,^5^ is central to this plan. SP aims to address people’s social, emotional, and practical needs, which are often closely related to their medical needs but not routinely addressed by clinical treatments.^6^ In the UK, SP typically consists of referral to a link worker, or other similar professional, who works with the individual to develop a personalised care plan that connects them to community support. This may include a wide range of activities such as sports, arts, volunteering, counselling, housing support, training, and employment advice.^7^

SP has been proposed as a way to reduce health inequalities by targeting the social determinants of health, including income, education, employment, housing, childhood experiences, and social support.^8^ Whilst SP may not alter these upstream determinants of health,^9,10^ it could improve access to and engagement with services, particularly among marginalised populations, thus reducing health inequalities as part of wider population health strategies.^11^ However, there is evidence that there are disparities in referrals to SP, with males, CYP under 16 years old, people from minority ethnic groups, and those facing more socioeconomic disadvantages less likely to be offered SP.^12–14^ There has been uneven embedding of SP link workers within services across the UK, with some indication that there are fewer link workers employed in areas with high health and social care needs (where they are needed most).^13^

Alongside geographical inequity in link worker posts, there is a “postcode lottery” in community assets, whereby more deprived areas generally have fewer resources available (e.g. groups, arts and cultural activities, leisure centres, sports clubs, libraries).^10,15–19^ This is a crucial consideration as SP relies on referring people to these existing resources. This dependency on local assets aligns with broader NHS policy shifts towards delivering care closer to people’s homes through neighbourhood-based models of care. Integrated neighbourhood approaches bring together professionals from primary care, community health services, social care, and the voluntary sector to coordinate holistic support within communities and proactively address health needs before they escalate. These models emphasise the importance of local infrastructure, community organisations, and accessible services in supporting prevention and wellbeing.^20^ Given the inequalities in provision, it has been suggested that SP may exacerbate health inequalities, rather than reducing them.^9,21^ Yet, more recent evidence indicates that progress is being made, with increasing representation of people living in more deprived areas, minority ethnic groups, and younger adults in referrals to SP between 2017 and 2023.^7,22^

Despite being funded as an “all-age” approach in the UK,^23^ SP research, policy, and practice have predominantly focused on adults.^12,24,25^ This is particularly problematic because adult SP approaches may not be suitable for CYP, who have different social contexts^26^ and face distinct challenges during adolescence, a period in which 75% of mental health problems emerge.^27^ Additionally, CYP and adults may access SP through differing routes. The typical delivery model in the UK (GP referral to a link worker based in primary care) was developed based on adults’ help-seeking behaviours. CYP, in contrast, do not always feel comfortable going to their GP for mental health or wellbeing support^28^ and may prefer to access SP through self-referral, educational institutions, and social care.^24^ However, it is unclear whether this is happening in practice.

Administrative data are uniquely placed to evaluate the implementation of SP as they capture real-world heterogeneity, allowing us to explore how CYP are currently accessing SP and whether there are inequalities in who receives SP. Link workers often use electronic health systems like EMIS or SystmOne, but many use bespoke systems designed for SP,^29^ including Elemental,^30^ Joy,^31^ and Social Rx Connect.^32^ These platforms allow healthcare professionals to refer people to SP and for SP practitioners (e.g. link workers, community navigators) to refer or signpost to community services.

Platforms can be used to monitor referrals, connect with services, record appointments and prescriptions, and measure impact. Additionally, platforms capture non-primary care referrals and provide rich data on SP interventions received, providing major advantages over GP records. Yet, although a growing body of research has used administrative records to evaluate SP among adults,^7,14,22,33–35^ to our knowledge only one study has focussed on CYP, which was limited to a small region of England.^36^

In this study, we aimed to use administrative data captured in a SP platform to describe the characteristics of CYP referred to SP across the UK and understand what SP looks like for these CYP, including the amount of support given and the number and types of interventions prescribed. Additionally, we aimed to explore whether access to and experiences of SP vary with age and have changed from 2017 to 2025. Overall, we hoped to identify whether SP reduces or exacerbates health inequalities among CYP, and whether this has changed over time.

## Methods

### Data

Access Elemental is a cloud-based SP platform used by health and social care professionals, community development workers, and other service providers to keep track of SP activities and their impact from the point of referral (https://www.theaccessgroup.com/en-gb/our-brands/elemental/). Access Elemental data are available since January 2017, and this study used data up to 30^th^ August 2025. During this period, Access Elemental captured data from 649,386 cases of SP among 508,449 individuals. For this analysis, we restricted data to individuals residing in the UK (0.5% cases excluded) who were aged between 4 and 25 years (90% cases excluded). This age range is in line with the group most often eligible for youth SP services.^24^ Children younger than 4 are likely too young to be able engage in SP, having not yet started compulsory education, meaning referrals may have captured a combination of infant and parental needs. Child and Adolescent Mental Health Services (CAMHS) offering extended or specialised care will support young people up to age 25. For CYP with more than one case of SP recorded, only the first case was retained (15% cases excluded). This left a final analytical sample size of 52,585 CYP, who were from over 700 SP sites across all regions of the UK. Analysis of these routine data was approved by UCL Research Ethics Committee (project 23909/002). Reporting follows the RECORD guidelines.^37^

### Measures

#### Individual characteristics

Access Elemental records basic demographic information, including age at time of referral (years), gender, and ethnicity. It categorised gender as female, male, non-binary, transgender, other, or not disclosed (not yet disclosed, prefer not to say). Ethnicity could not be used due to the high proportion of missing data (83% missing; Table S1), as in previous analyses of Access Elemental and other SP records.^7,33,36^

We determined country of residence (England, Wales, Scotland, Northern Ireland) from individuals’ postcodes. Where postcodes were missing (24%), we used the SP site location. Postcodes were also used to measure area urbanicity and index of multiple deprivation (IMD) via postcode linkage to the Office for National Statistics Postcode Directory.^38^ Urbanicity (urban, rural) was derived for the Output Area (OA) in which CYP lived using the 2001 census urban/rural indicators, with definitions of urban vs rural allowed to differ across countries.^38^ IMD was calculated for the Lower Layer Super Output Areas (LSOA) in which the individual lived and represented the rank of that LSOA within the country. We categorised the IMD ranks into deciles within each country, from the most deprived (1) to the least deprived (10).^38^ In the general population, roughly 10% of people live in each deprivation decile in each country.

The source of each referral into SP was recorded as medical vs non-medical, with no further detail available from Access Elemental. Medical referrals were mainly from GPs but could also include allied health workers such as occupational therapists, physiotherapists, discharge teams, and hospital-based social workers. Non-medical referrals were from educational institutions, local authorities, community organisations, self-referrals, and housing associations, among others.

Over 600 reasons for referral were recorded, which we grouped into seven domains.^7^ Grouping was done independently by two authors with disagreements discussed and resolved by consensus, in line with previous analyses of Access Elemental data.^7^ Multiple referral reasons were allowed across:

1. Mental health, psychological, or behavioural needs
2. Physical health and wellbeing
3. Social relationships (e.g. loneliness, social isolation)
4. Lifestyle (e.g. exercise, diet, substance use)
5. Employment, education, and skills
6. Practical support (e.g. housing, finances)
7. Other reasons

#### Social prescribing pathway

Link workers recorded the date of their first contact with the client, which could be an introductory appointment, text, or call. Subsequent contacts could include face-to-face or online meetings, visiting community venues together, calls, texts, emails, or letters, among other forms of contact. We summed the total number of contacts and used the first contact date to indicate the year in which SP was delivered. Where contact type was reported (n=30,394), we categorised it into eight categories (face to face, phone call, text, online/video call, email, letter, other, unspecified). Where complete information was available on contact time (n=17,328), we also reported the average length of each contact and estimated the average amount of time spent by the link worker on each case by summing the time spent in all contacts. Case status was recorded as complete, active, waiting, or other.

Contacts may lead to individuals being connected with community resources, either through onward referrals, which we hereby refer to as being prescribed an intervention, or through signposting. We report both the number of signposts and number of interventions recorded by the link worker. These are assessed separately as distinct outcomes, with signposting likely to involve less interaction with link workers than being prescribed an intervention. However, an absence of recorded signposts/interventions does not necessarily mean that none were made. Additionally, a recorded signpost/intervention indicates only that it was issued by the link worker but does not imply subsequent attendance or engagement. In total, over 400 intervention types were recorded. Following the procedure used for referral reasons above, we classified interventions into the same seven domains, which were not mutually exclusive. Intervention format was also recorded, which we classified as an event, programme, service, course, or other. We could not examine the domain or format of signposts because this information was missing for 77% of individuals.

### Statistical analysis

We first summarised the characteristics of CYP referred to SP, described the SP pathway, and explored intervention domains and formats. We then evaluated whether all individual characteristics and the SP pathway differed according to age, using a series of unadjusted regression models to test their associations with age (measured continuously in years). We used logistic regression for binary outcomes, multinomial logistic regression for categorical outcomes, and negative binomial regression for count outcomes (due to overdispersion). All regression models included cluster-robust standard errors to account for clustering of individuals within SP sites. To plot differences according to age, we classified participants into age groups following the key stages of UK educational provision: 4-7, 8-11, 12-14, 15-18, 19-21, and 22-25 years. Finally, to explore how the characteristics of CYP referred to SP have changed over time, we used separate regression models to test the associations between the year in which SP was delivered and all individual characteristics, specifically age (linear regression), gender (multinomial logistic regression), referral source (logistic regression), and referral reasons (logistic regression). Missing data were handled using listwise deletion separately for each analysis due to the lack of auxiliary variables to enable multiple imputation.

### Patient and public involvement

The design of this study was informed by roundtables with young people, social prescribing practitioners, and policymakers organised through the Social Prescribing Youth Network. Patients and public were not involved at subsequent stages of the analysis or write up.

## Results

### Individual characteristics

A total of 52,585 individuals aged 4 to 25 years were referred to SP services using Access Elemental in the UK between January 2017 and August 2025. Among these CYP, the average age was 20.04 years (standard deviation [SD]=4.71; Figure 1A), 57% were female, 39% male, <2% were in other gender groups, and 3% did not disclose their gender (Figure 1B). The large majority of CYP accessing SP were from England (79%), with 11% in Wales, 7% in Northern Ireland, and 3% in Scotland (Table S2). Within England, the majority of CYP resided in the North West (36%), London (18%), and the North East (11%). The least represented regions of England were the West Midlands (0.4%) and the East Midlands (0.4%; Figure S1). Although the regional distribution of CYP within England was similar to the distribution of all Access Elemental cases (i.e. all age groups; Figure S1), CYP were over-represented in Wales and Northern Ireland compared to all cases.

**Figure 1.**
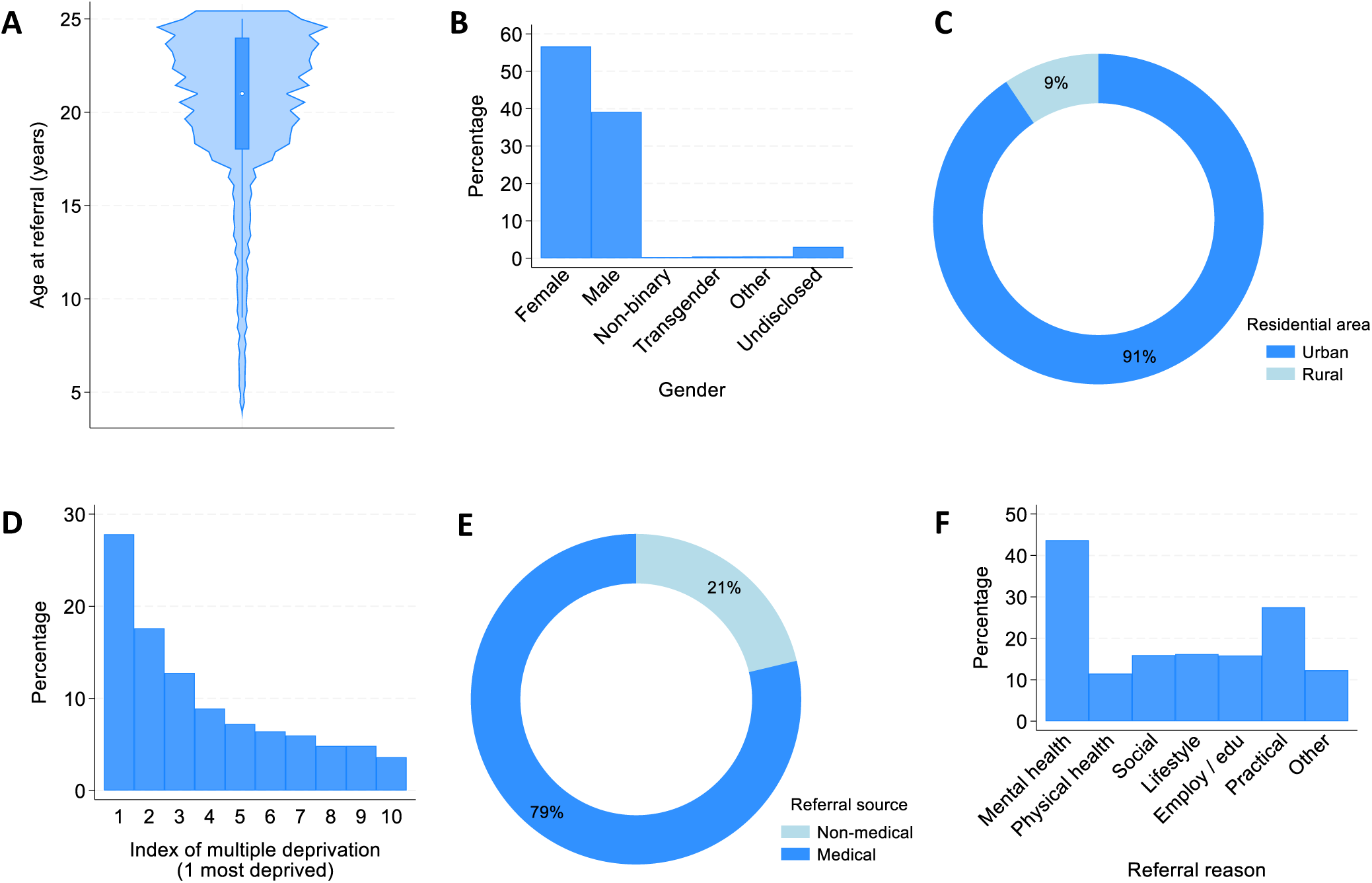
Descriptive statistics showing the characteristics of individuals referred to social prescribing: A) age in years at referral (n= 52,585), B) gender (n= 52,585), C) residential area (n= 52,585), D) neighbourhood index of multiple deprivation (n= 52,585), E) referral from a non-medical source, including educational institutions, local authorities, community organisations, self-referrals, and housing associations, or a medical source, including GPs, occupational therapists, physiotherapists, discharge teams, and hospital-based social workers (n= 52,585), and F) referral reason, in which each individual could have multiple referral reasons (n= 52,585).

Most CYP lived in urban areas (91%; Figure 1C). SP users were more likely to be from the most deprived areas, as 28% lived in the most deprived IMD decile, 18% in the second decile, and 13% in the third decile. In total, 58% of CYP referred to SP lived in the top three most deprived areas. Only 4% of CYP were living in the least deprived IMD decile (Figure 1D). In comparison to all Access Elemental cases (i.e. all age groups), CYP were more likely to be in the lowest IMD deciles (Figure S2).

A large majority of CYP (79%) were referred to SP via medical routes (Figure 1E). The most common reason for referral was mental health psychological or behavioural needs (44%), followed by practical support (27%), and social relationships, lifestyle, or employment education and skills (all 16%; Figure 1F). The least common referral reason was physical health and wellbeing (11%). Most CYP (70%) had only one referral reason recorded. Among those referred for mental health psychological or behavioural needs, 52% had another referral reason, primarily social relationships (21%), practical support (21%), or employment education and skills (12%).

### Social prescribing pathway

The number of contacts related to each SP case ranged from 0 to 357, with a mean of 4.64 (SD=7.70, median=2). Overall, 88% of CYP had at least 1 contact with a link worker, and 77% had between 1 and 10 contacts. Looking at SP case status, 77% of cases were closed, 14% active, and 9% were waiting for SP. As expected, the average number of contacts was higher among closed cases than among active cases or those waiting for SP (Figure 2A). Across all recorded contacts, phone calls were most common (44%), followed by texts (22%), and emails (13%). Face to face appointments comprised 8% of contacts (Figure 2C). The most common contact length was 0-15 minutes (66% of contacts; Figure 2D). In total, for cases with complete data on contact length (n=17,328), link workers spent a mean of 66 minutes (SD=108 minutes, median=38 minutes) on each case, but durations varied from 0 minutes to 51 hours. Link workers had spent more time with closed and active cases, than with those waiting for SP (Figure 2B).

**Figure 2.**
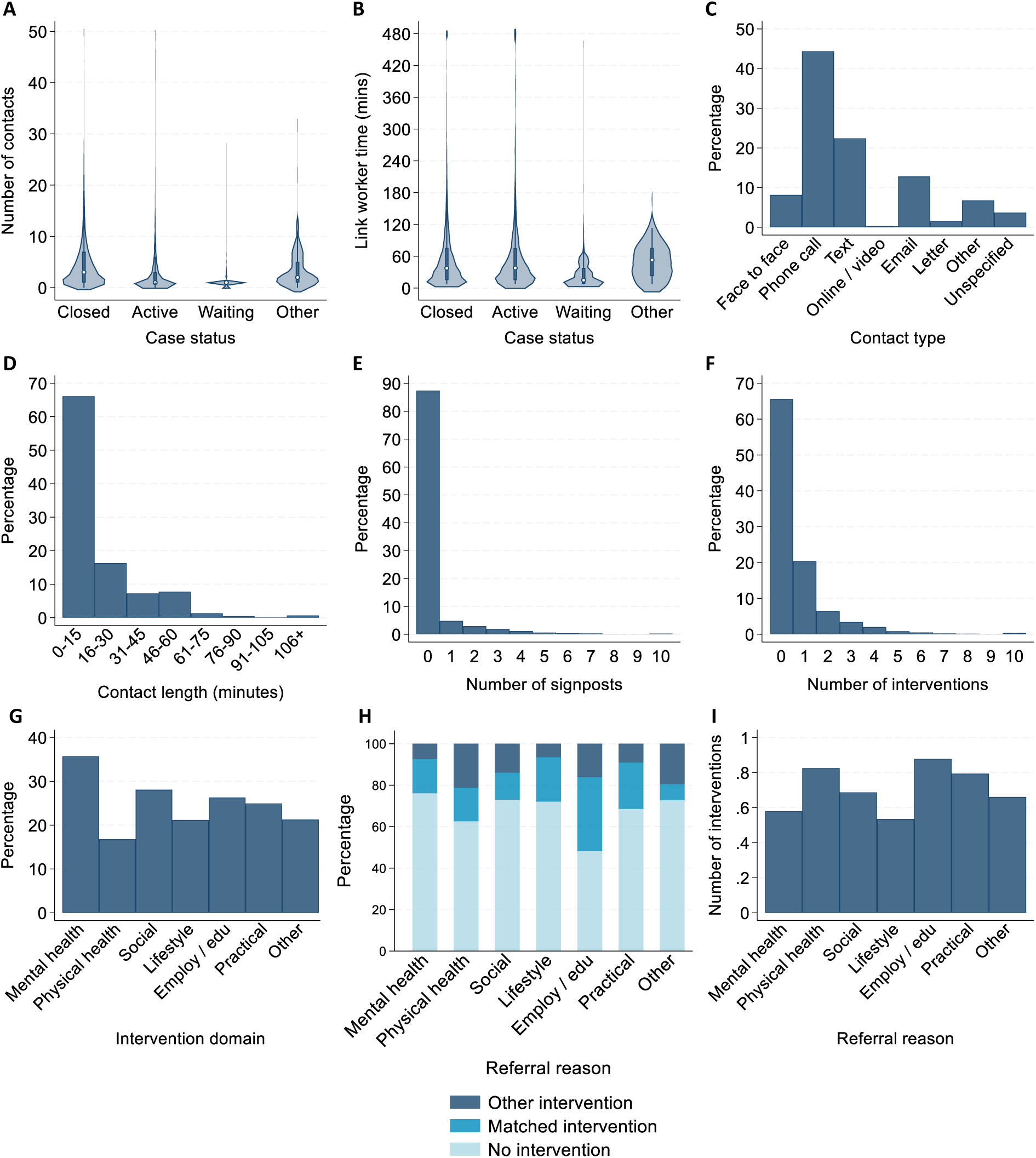
Descriptive statistics showing the details of the social prescribing received by children and young people: A) number of contacts according to case status, winsorized at 50 contacts for clarity (n=52,585), B) total amount of time spent by link workers on each case according to case status, winsorized at 480 minutes (8 hours) for clarity (n=17,328), C) proportion of recorded contact types (n=39,065), D) average contact length in minutes for each contact (n=32,354), E) total number of recorded signposts, winsorized at 10 for clarity (n=52,585), F) total number of recorded interventions, winsorized at 10 for clarity (n=52,585), G) proportion of recorded interventions in each domain (n=15,990), H) proportion of cases receiving an intervention matched to referral reason in each referral reason domain, in which “other intervention” indicates they did not receive an intervention in a domain matched to their referral reason, but did get one or more other interventions (n=50,326), and I) total number of interventions recorded according to referral reasons (n=52,423).

Looking at onward signposting, 13% had one or more signposts recorded (Figure 2E). Among them, the number of signposts ranged from 1 to 47, with a mean of 2.71 (SD=2.50, median=2). Focussing instead on prescriptions, 34% of CYP had one or more interventions recorded (Figure 2F). Among them, the number of interventions ranged from 1 to 28, with a mean of 1.94 (SD=1.77, median=1). The most common intervention format was services (61%), followed by programmes (23%), and courses (9%).

Events (2%) and other intervention types (5%) were less common. Among cases with intervention domains recorded, 36% were given an intervention for mental health psychological or behavioural needs, followed by 28% for social relationships, 26% for employment education and skills, and 25% for practical support (Figure 2G). The least common intervention domain was physical health and wellbeing (17%).

The proportion of CYP who received an intervention matching their referral reasons varied across domains (Figure 2H). Those referred for employment education and skills were most likely to have an intervention recorded (52%), and to have an intervention matched to referral reasons (36%). Those referred for mental health were least likely to have an intervention recorded (24%), but 17% received an intervention in the mental health domain. Apart from ‘other’ referral reasons, those referred for social relationships were least likely to receive a matched intervention, as only 13% of social relationship referrals had a social relationship intervention recorded.

### Differences by age

In this sample of CYP aged 4 to 25 years, older individuals were less likely to be male than female (relative risk ratio [RRR]=0.97, 95% confidence interval [CI]=0.96, 0.97, p<0.001; Figure 3A). Although descriptive statistics indicated that older individuals were more likely to be referred to SP via medical routes (Figure 3B), there was no evidence for a linear association between age and odds of medical referral (Table 1). Looking at referral reasons (Figure S3), increasing age was associated with higher odds of being referred for mental health psychological or behavioural needs (odds ratio [OR]=1.04, 95% CI=1.00, 1.09, p=0.033), social relationships (OR=1.05, 95% CI=1.02, 1.07, p=0.001), practical support (OR=1.08, 95% CI=1.03, 1.14, p=0.002), and, in contrast, lower odds of being referred for physical health and wellbeing (OR=0.91, 95% CI=0.83, 0.99).

**Figure 3.**
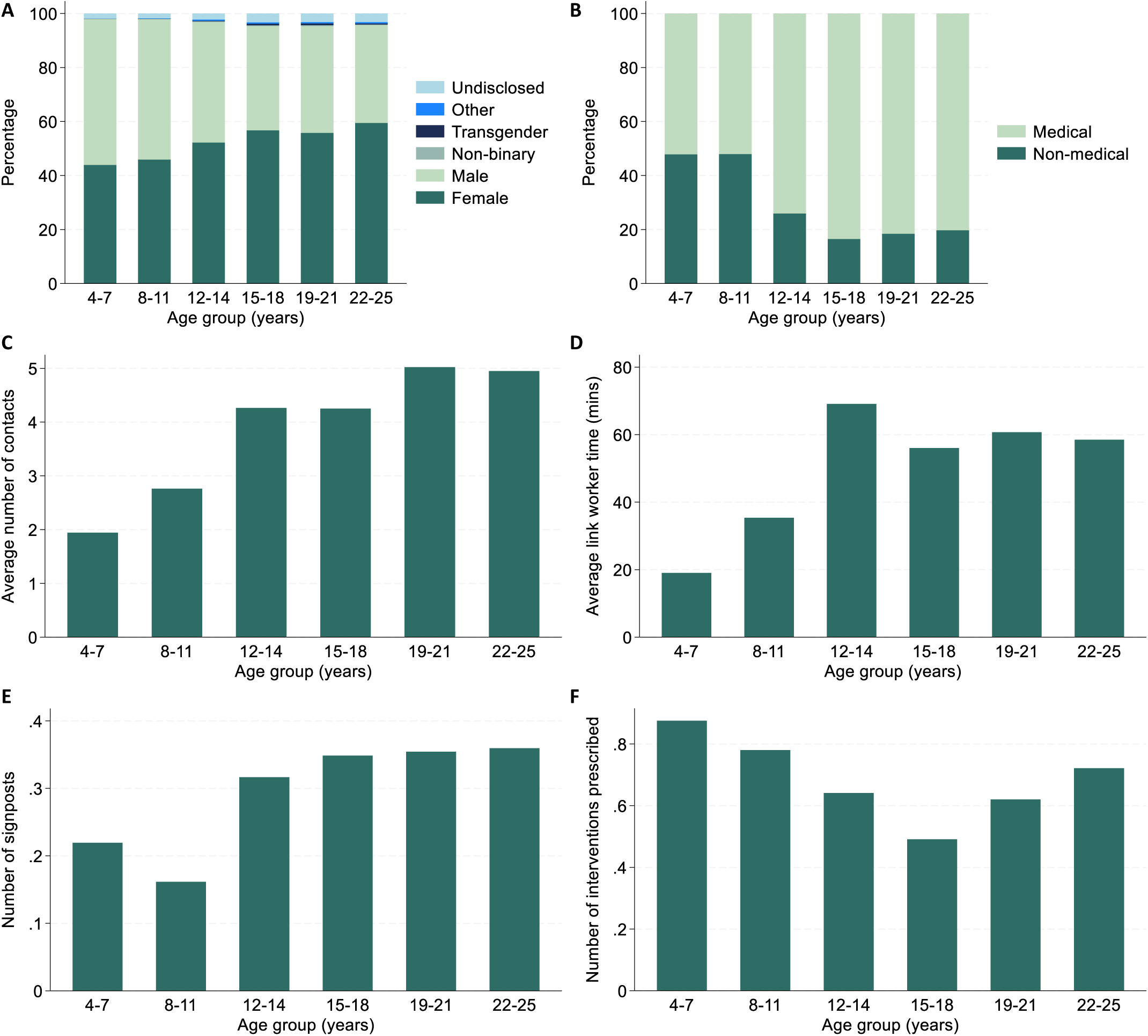
Descriptive statistics showing the difference in the following individual characteristics and social prescribing pathway across age groups (4-7, 8-11, 12-14, 15-18, 19-21, 22-25 years): A) gender (n=52,585), B) referral source (n=47,069), C) number of link worker contacts (n=52,585), D) link worker time spent on the case (n=17,328), E) number of signposts (n=52,585), and F) number of interventions prescribed (n=52,585). See Figure S2 for differences in referral reasons across age groups.

**Table 1.** Results from regression models testing the associations between age (in years), individual characteristics, and characteristics of the social prescribing pathway received.

|  | Model type | N | Coef | 95% CI | p value |
| --- | --- | --- | --- | --- | --- |
| **Gender** (ref: female) | Multinomial logistic (RRR) | 52,585 |  |  |  |
| Male |  |  | **0.96** | **0.96, 0.97** | **<0.001** |
| Non-binary |  |  | 1.02 | 0.98, 1.05 | 0.307 |
| Transgender |  |  | 1.01 | 0.98, 1.04 | 0.444 |
| Other |  |  | 1.03 | 0.99, 1.07 | 0.090 |
| Not disclosed |  |  | 1.01 | 0.98, 1.05 | 0.501 |
| **Medical referral source** | Logistic (OR) | 47,069 | 1.07 | 0.97, 1.17 | 0.186 |
| **Referral reason** |  |  |  |  |  |
| Mental health | Logistic (OR) | 52,423 | **1.04** | **1.00, 1.09** | **0.033** |
| Physical health/wellbeing | Logistic (OR) | 52,423 | **0.91** | **0.83, 0.99** | **0.031** |
| Social relationships | Logistic (OR) | 52,423 | **1.05** | **1.02, 1.07** | **0.001** |
| Lifestyle | Logistic (OR) | 52,423 | 0.98 | 0.91, 1.05 | 0.588 |
| Employ/education/skills | Logistic (OR) | 52,423 | 1.03 | 0.99, 1.07 | 0.106 |
| Practical support | Logistic (OR) | 52,423 | **1.08** | **1.03, 1.14** | **0.002** |
| Other reasons | Logistic (OR) | 52,423 | 0.96 | 0.92, 1.01 | 0.086 |
| **Link worker contacts** | Negative binomial (IRR) | 52,585 | **1.04** | **1.01, 1.07** | **0.009** |
| **Link worker time** | Linear (B) | 17,328 | -0.08 | -2.15, 1.99 | 0.937 |
| **Number of signposts** | Negative binomial (IRR) | 52,585 | 1.03 | 0.96, 1.10 | 0.444 |
| **Number of interventions** | Negative binomial (IRR) | 52,585 | 1.00 | 0.96, 1.05 | 0.860 |
| **Intervention domain** |  |  |  |  |  |
| Mental health | Logistic (OR) | 15,990 | 1.01 | 0.91, 1.11 | 0.851 |
| Physical health/wellbeing | Logistic (OR) | 15,990 | 0.94 | 0.88, 1.02 | 0.128 |
| Social relationships | Logistic (OR) | 15,990 | **0.94** | **0.89, 0.99** | **0.020** |
| Lifestyle | Logistic (OR) | 15,990 | 0.98 | 0.88, 1.10 | 0.738 |
| Employ/education/skills | Logistic (OR) | 15,990 | **1.07** | **1.03, 1.12** | **0.001** |
| Practical support | Logistic (OR) | 15,990 | **1.23** | **1.13, 1.35** | **<0.001** |
| Other reasons | Logistic (OR) | 15,990 | 0.93 | 0.83, 1.05 | 0.245 |

Note. Bold text indicates $p < 0.05$ .

Within the SP pathway, older individuals had more link worker contacts (incidence rate ratio=1.04, 95% CI=1.01, 1.07, p=0.009). However, there was no evidence for associations with link worker time or number of interventions or signposts recorded. Examining intervention domains separately, older individuals had higher odds of being prescribed interventions for employment education and skills (OR=1.07, 95% CI=1.03, 1.12, p=0.001) and practical support (OR=1.23, 95% CI=1.13, 1.35, p<0.001), but lower odds of interventions for social relationships (OR=0.94, 95% CI=0.89, 0.99, p=0.020).

### Changes over time

Finally, we tested whether the characteristics of CYP referred to SP have changed over time. There was evidence that the average age of those referred to SP has increased from 2017 to 2025 (coef=0.29, 95% CI=0.01, 0.58, p=0.045; Figure 4A). Compared to females, the likelihood of individuals identifying as non-binary or transgender or not disclosing their gender all increased over time (Figure 4B, Table 2).

**Figure 4.**
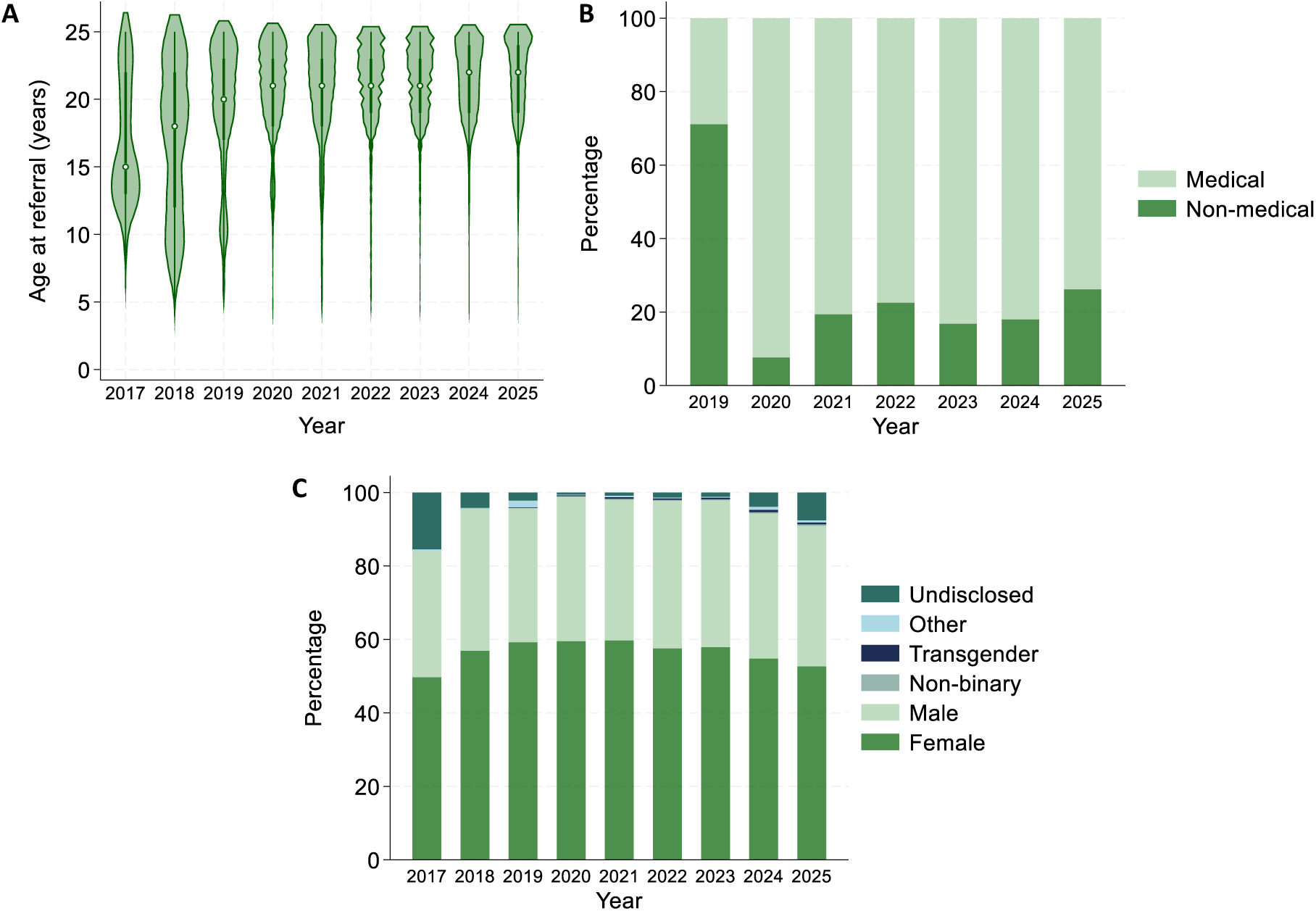
Descriptive statistics showing the difference in individual characteristics according to the year in which individuals received social prescribing: A) age (n=46,227), B) referral source (n=41,231), C) gender (n=46,227). See Figure S3 for changes in referral reasons over time.

**Table 2.**
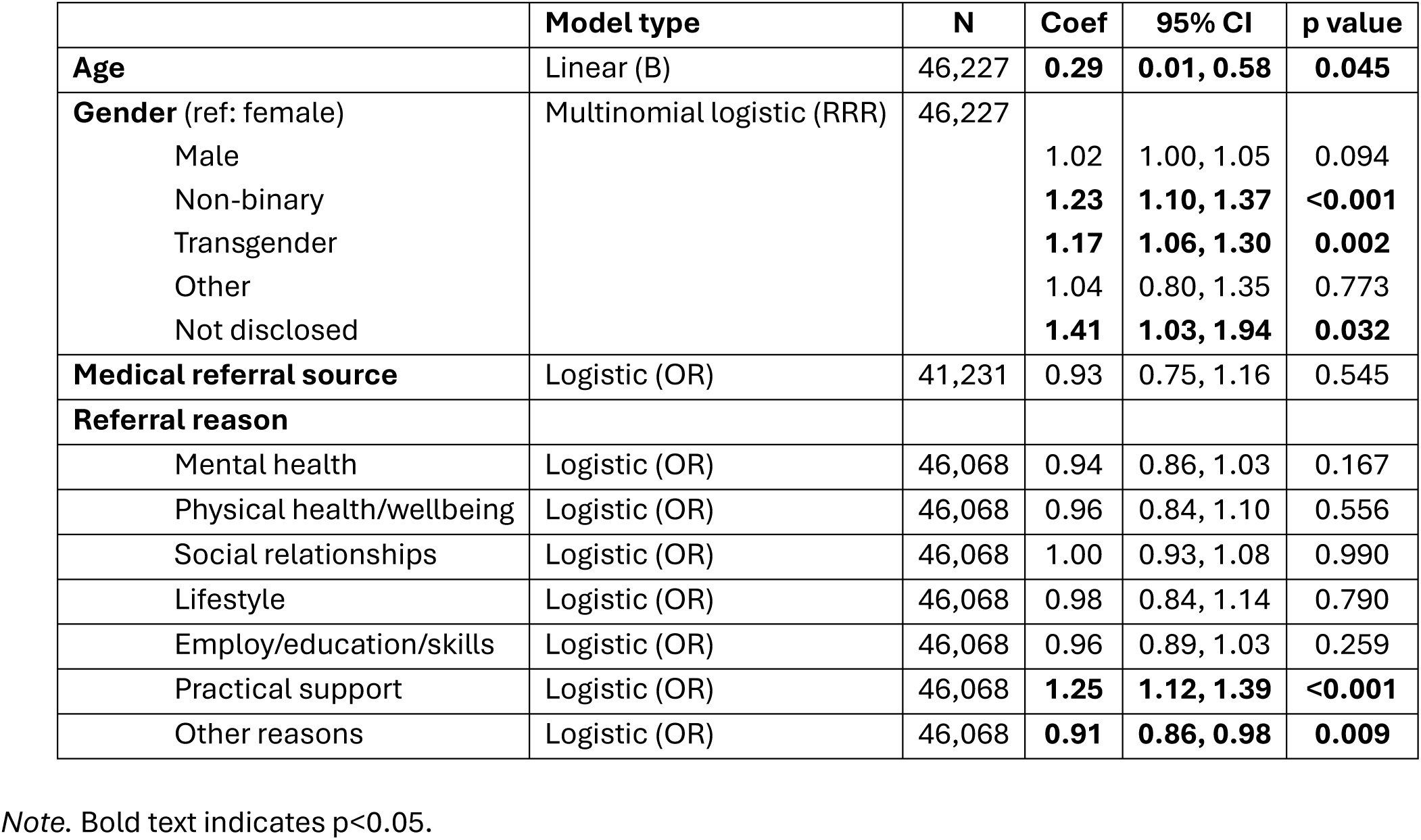
Results from regression models testing the associations between year in which social prescribing was received and individual characteristics.

|  | Model type | N | Coef | 95% CI | p value |
| --- | --- | --- | --- | --- | --- |
| **Age** | Linear (B) | 46,227 | **0.29** | **0.01, 0.58** | **0.045** |
| **Gender** (ref: female) | Multinomial logistic (RRR) | 46,227 |  |  |  |
| Male |  |  | 1.02 | 1.00, 1.05 | 0.094 |
| Non-binary |  |  | **1.23** | **1.10, 1.37** | **<0.001** |
| Transgender |  |  | **1.17** | **1.06, 1.30** | **0.002** |
| Other |  |  | 1.04 | 0.80, 1.35 | 0.773 |
| Not disclosed |  |  | **1.41** | **1.03, 1.94** | **0.032** |
| **Medical referral source** | Logistic (OR) | 41,231 | 0.93 | 0.75, 1.16 | 0.545 |
| **Referral reason** |  |  |  |  |  |
| Mental health | Logistic (OR) | 46,068 | 0.94 | 0.86, 1.03 | 0.167 |
| Physical health/wellbeing | Logistic (OR) | 46,068 | 0.96 | 0.84, 1.10 | 0.556 |
| Social relationships | Logistic (OR) | 46,068 | 1.00 | 0.93, 1.08 | 0.990 |
| Lifestyle | Logistic (OR) | 46,068 | 0.98 | 0.84, 1.14 | 0.790 |
| Employ/education/skills | Logistic (OR) | 46,068 | 0.96 | 0.89, 1.03 | 0.259 |
| Practical support | Logistic (OR) | 46,068 | **1.25** | **1.12, 1.39** | **<0.001** |
| Other reasons | Logistic (OR) | 46,068 | **0.91** | **0.86, 0.98** | **0.009** |

Note. Bold text indicates $p < 0.05$ .

Referral source was only recorded from 2019 onwards, although it was missing for most individuals until 2021. Therefore, although Figure 4C shows early changes in the proportion of individuals referred from medical sources, this was likely skewed by missing data in 2019-2020. In a logistic regression model, there was no evidence for an association between year and referral source. For referral reasons (Figure S4), there was only evidence that the odds of being referred for practical support increased from 2017 to 2025 (OR=1.25, 95% CI=1.12, 1.39, p<0.001) and the odds of being referred for other reasons decreased over time (OR=0.91, 95% CI=0.65, 0.98, p=0.009).

## Discussion

This is the first study to explore access to and delivery of SP for CYP across the UK. In total, 52,585 CYP were referred to SP services using the Access Elemental platform (one of several bespoke electronic health systems for SP) from 2017 to August 2025. The majority of CYP were aged 18 and over, female, and lived in more deprived urban areas. Most were referred by GPs or other allied health workers, and mental health was the leading reason for referral. The care received varied widely, with the typical pathway including five link worker contacts totalling just over an hour. Around a third received an onward referral to community support, most commonly an intervention for mental health or social relationships. There were only minor differences in the characteristics of those referred to SP and the SP pathway by age, but the average age of those referred to SP increased over time.

Our first aim was to describe the characteristics of those referred to SP in the UK. The gender disparity in referrals was very similar to findings from adults, which have shown that almost twice as many women access SP than men.^12^ Additionally, given that SP research, policy, and practice have predominantly focused on adults,^12,24,25^ it was unsurprising that CYP under 18 were underrepresented in our sample. This could partly be driven by using data from one electronic health record system, as adult SP services may be more likely to use Access Elemental than child and adolescent services. However, when considered alongside previous findings that people under 30 represent a very small proportion of those referred to SP across England,^22^ and evidence for under-referring of under 15s to SP in North West England (and 15-29-year-olds to a lesser extent),^14^ our findings suggest that SP is not reaching all children and adolescents who could benefit. The large proportion of CYP from England, compared to other UK nations, may have been driven by the distribution of services that use Access Elemental.

However, CYP were more likely to live in Wales or Northern Ireland than adults who were referred to SP via Access Elemental. This may reflect varying approaches to SP in devolved nations, as differences in link worker employment models, funding, and integration with primary care could all influence access to CYP SP services.^39,40^

Given previous evidence that adults with lower socioeconomic position were less likely to be offered SP, and fewer link workers were employed in more deprived areas,^12–14^ it was somewhat surprising that CYP were more likely to be from the most deprived areas, with over a quarter living in the most deprived IMD decile. Together with evidence for increasing referrals to SP in more deprived areas from 2017 to 2023,^7,22^ this suggests that SP is already operating in a way that could reduce health inequalities and reaching the populations that it should. Several factors could be driving the higher referrals to SP among more deprived areas, including greater need, recent changes in link worker provision (since previous research),^13^ or a higher proportion of services using Access Elemental. Whatever the underlying causes, this shows the importance of avoiding accidental “decommissioning” of SP, a concern that has arisen since the Additional Roles Reimbursement Scheme (which funds link workers within primary care networks) has broadened, leading to reductions in link workers and link worker hours. Our findings indicate that thousands of CYP would miss out if SP stopped being offered in the UK, particularly those in the most deprived areas.

We also expected the proportion of CYP referred from non-medical routes (e.g. educational institutions, local authorities, community organisations, self-referrals, and housing associations) to be higher than among adults. Instead, we found that 79% of CYP were referred via medical routes (GPs or other allied health workers). This was very similar to referral patterns in a small evaluation of one CYP SP service that used the Joy platform (in which 74% medical referral routes),^36^ as well as patterns in the general population (83% had medical referral routes).^33^ Although this suggests that CYP are not more likely to be referred to SP from alternative routes, as previously found,^24^ it only reveals current patterns. Our dataset just included CYP who had been referred to SP, which is unlikely to be representative of all CYP who could benefit from SP. It is likely that if non-medical referral routes were more easily accessible and more widely promoted, even more CYP might access SP. Our findings could also be partially due to the sporadic use of Access Elemental across services, so should be confirmed using more data from other SP platforms.

Our second aim was to understand what SP looks like for CYP. On average, individuals had less than 5 contacts (of any type) with a link worker, lasting around an hour in total. This was much lower than the six contact hours received on average in a previous evaluation of one English CYP SP service, likely because the earlier evaluation focussed on CYP successfully discharged from SP.^36^ However, our current findings are similar to the wider population referred to SP via Access Elemental,^7^ suggesting that SP is often not delivered in line with best practice guidelines, which recommend 6-12 contacts over a three-month period.^41,42^ Additionally, only a third had an onward referral to an intervention recorded, so SP often may not result in connection to community support. It is unclear whether this is driven by the limited time spent with link workers, individual preferences, a lack of local community resources, incomplete recording of interventions, or a combination of these factors. Consistent with previous evidence among CYP,^36^ the most common reason for referral was mental health psychological or behavioural needs (recorded for 44%). Yet CYP referred for this reason were least likely to have an intervention recorded, potentially indicating a lack of appropriate community support on offer for this group. Disentangling why rates of onward referrals were so low should be a key priority for future research.

Our final aims were to explore whether access to and experiences of SP varied with age and over time. We defined the target population broadly, including CYP aged 4 to 25 years. It is worth considering that SP may be very different for individuals at either end of this age range. The youngest are completely reliant on parents/carers and probably involved in parent or family SP, as they cannot engage in the “what matters to you” conversation that is a crucial component of individual-focussed SP.^42^ There were relatively few differences in the characteristics of those referred to SP across age or time, but we did find that the average age of CYP referred to SP has increased from 2017 to 2025. This is concerning, as it appears that the bias towards adults in SP services is getting worse, despite increasing numbers of link workers being employed during this period.^43^ Further research is vital to understand this finding so that provision of and access to CYP SP can be improved across the country.

We also found that older CYP were more likely to be prescribed interventions for employment education and skills, although there were no age differences in being referred for this reason. In Europe, SP has been specifically developed for young people Not in Employment, Education, or Training (NEET).^44,45^ Our findings suggest that this could already be happening in practice for young adults in the UK, and practice could be developed further support NEET youth. Additionally, older CYP were less likely to be prescribed interventions for social relationships and more likely to be prescribed interventions for practical support. This is likely to reflect people’s changing needs during adolescence, as the earlier years are more focussed on establishing strong peer relationships and sense of self,^46,47^ before adolescents then start to become independent and are thus likely to need more practical support with tasks like finances, housing, utilities, and food. SP practitioners should be aware of these developmental trends, as well as using them to support in service planning and provision.

This study had several strengths, such as the use of data from the most widely adopted SP platform in the UK, including a large and diverse population of CYP. For the first time, we were able to provide detailed information on which CYP are currently being referred to SP across the UK, and what SP looks like for them, as well as differences across age groups and the eight-year period. Access Elemental includes CYP referred to SP through medical and non-medical pathways, which is key for understanding inequalities in service provision. However, administrative data are limited by missingness and varying data quality. For example, we could not explore the role of ethnicity, as it was missing for the large majority of CYP. It was unclear whether individuals with no recorded signposts/interventions did not receive any prescriptions or were missing these data. Additionally, we could not investigate referral source in further detail than medical vs non-medical routes, which remains a key priority for CYP SP. Enhancing recording and data quality thus remain key priorities for SP practitioners and platform providers, as previously identified.^7,14,36,48^

Furthermore, due to the nature of our data, we could only explore access to and delivery of SP by sites that have chosen to use the Access Elemental platform. We could not test whether residential characteristics of referred CYP or SP pathways differed over time, as these were considered likely to result from differences in the SP sites who chose (or could afford) to use Access Elemental over time. Relatively few SP sites were consistently included in this dataset from 2017 to 2025. It was also unclear whether evidence for changes in CYP characteristics over time (e.g. increasing age) were driven by these changes in SP sites. Testing how CYP SP has changed during this period, independent of the platform used to deliver SP, should be a focus of future research. Nevertheless, our findings were generally in line with a previous evaluation of CYP SP that used data from the Joy platform,^36^ indicating that they were not solely driven by the use of data from Access Elemental.

Overall, we aimed to identify whether SP has the potential to address health inequalities, or is more likely to be exacerbating inequalities, and whether this has changed over time. Using data from one electronic health record system, we found that SP currently has limited reach for CYP, particularly those under 18, and is not widely accessed through non-medical routes. Crucially, SP may have the potential to reduce health inequalities, as CYP referred to SP were more likely to live in deprived areas. This suggests that any reductions in number of link workers or link worker hours in England could have the largest negative impacts on the CYP most in need of support. However, among those referred to SP, there was wide variability in care pathways, with relatively few CYP connected with community support. It is possible that this finding was driven by limited recording and poor data quality, meaning understanding the delivery of CYP SP across the UK remains a priority, as well as providing robust evidence for its on impacts of health and wellbeing.

## Declarations

### Author contributions

JKB conceptualised the study and developed the analytical plan, with support from FB and DF. JKB performed analyses, with input from all other authors, and drafted the manuscript. All co-authors contributed to the interpretation and reporting of findings and read and approved the final version of the manuscript.

### Data sharing

Access Elemental data cannot be shared publicly. Code for the analyses is available from JKB on request.

## Acknowledgement

We thank Access Elemental for granting us data access and for the numerous conversations and input that enabled this evaluation.

## Supplementary Materials

**Figure S1.**
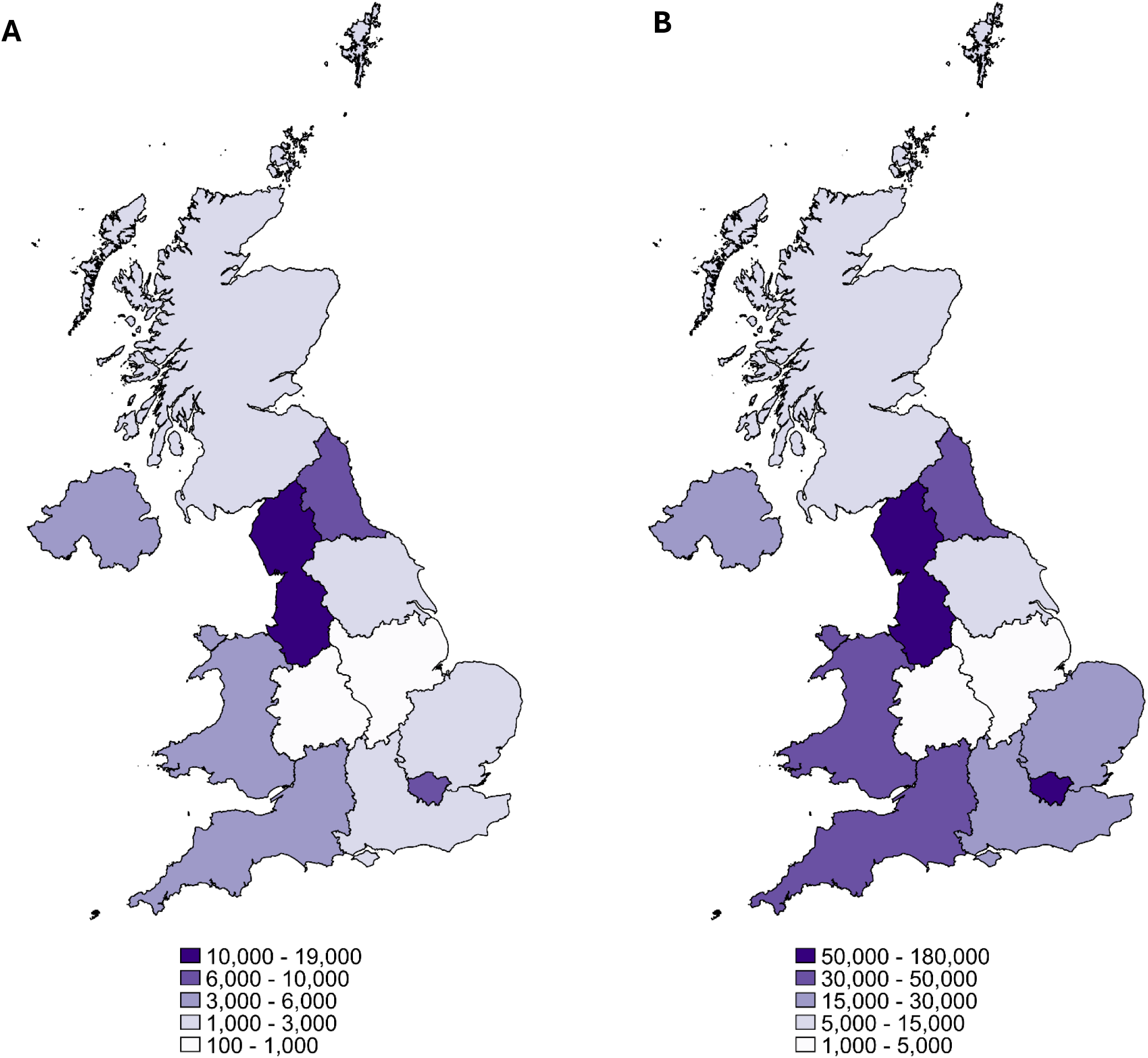
Maps showing A) the distribution of children and young people included in the analytical sample, compared to B) the distribution of all individuals (of any age) included in the Access Elemental dataset.

**Figure S2.**
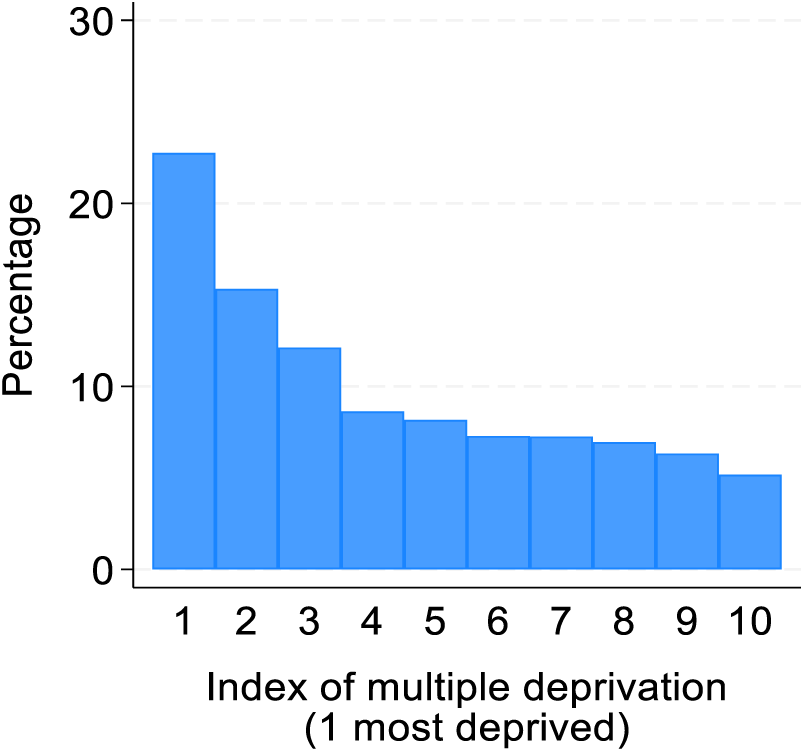
Descriptive statistics showing neighbourhood index of multiple deprivation for all individuals (of any age) included in the Access Elemental dataset (n=395,762).

**Figure S3.**
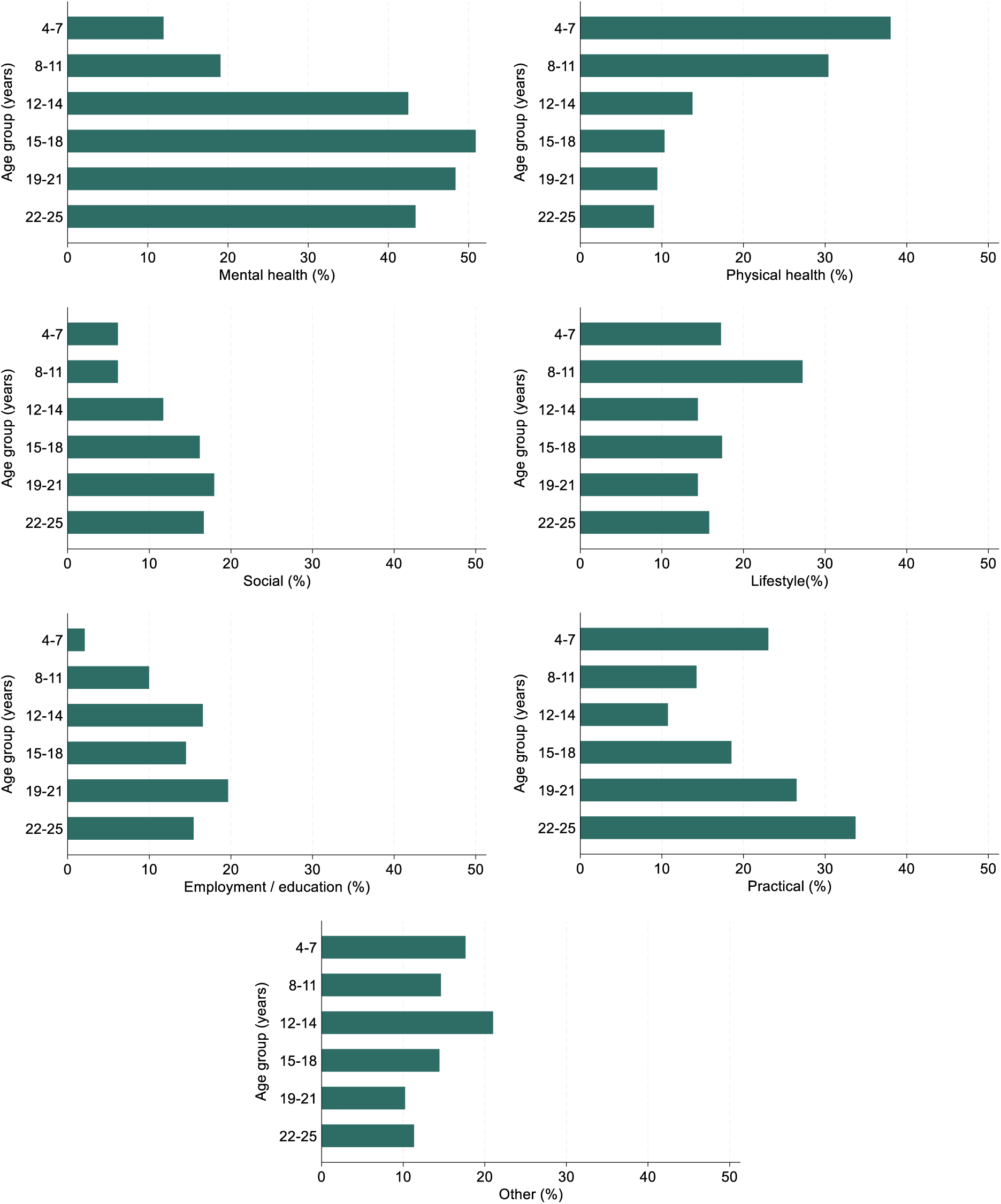
Descriptive statistics showing the difference in referral reasons according to age group (n=52,423). Each individual case may have multiple referral reasons.

**Figure S4.**
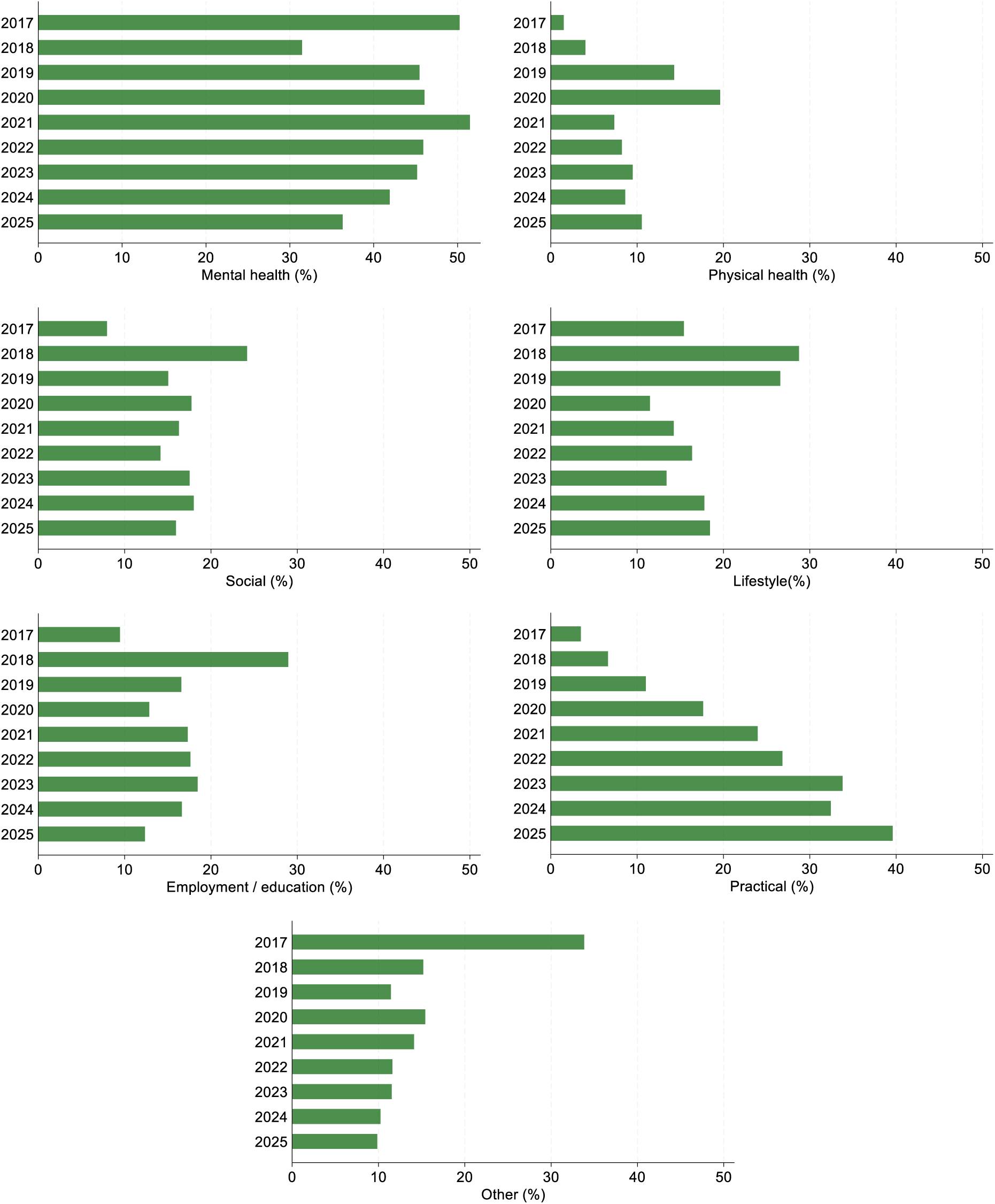
Descriptive statistics showing the difference in referral reasons according to the year in which individuals received social prescribing (n=46,068). Each individual case may have multiple referral reasons.

**Table S1.** Number of children and young people and proportion of sample (n=52,585) with missing data.

| Characteristic | Missing |
| --- | --- |
| Age | - |
| Gender | - |
| Ethnicity | 43,555 (83%) |
| Country | - |
| Urbanicity | 12,688 (24%) |
| Index of multiple deprivation (IMD) | 12,688 (24%) |
| Referral source | 5,516 (10%) |
| Referral reason | 162 (<1%) |
| Case status | - |
| Number of contacts | - |
| Year social prescribing delivered | 6,358 (12%) |
| Contact type<sup>a</sup> | 22,191 (42%) |
| Contact length<sup>a</sup> | 35,257 (67%) |
| Link worker time<sup>a</sup> | 35,257 (67%) |
| Number of signposts | - |
| Number of interventions | - |
| Intervention format | - |
| Intervention domain<sup>b</sup> | 2,104 (12%) |

<sup>a</sup> Includes those with no contacts recorded (n=6,357) as well as those with contacts recorded but this information missing (n=15,834 for contact type; n=28,900 for contact length/link worker time).
<sup>b</sup> Percentage of individuals with one or more interventions recorded (n=18,094), i.e. for whom these data should not be missing.

**Table S2.**
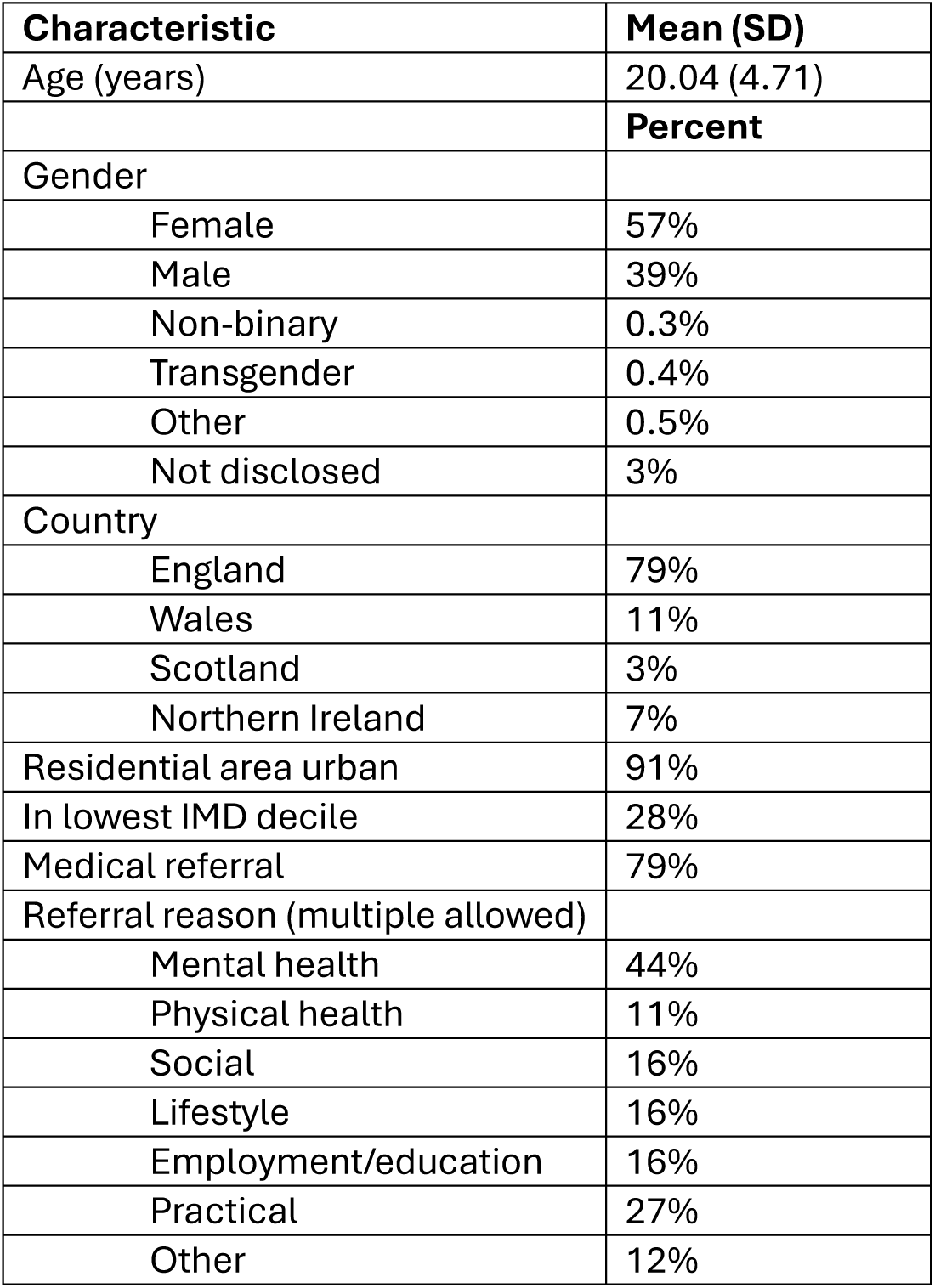
Characteristics of children and young people referred to social prescribing.

## References

1. Watt, T., et al. Health in 2040: Projected Patterns of Illness in England. https://www.health.org.uk/reports-and-analysis/reports/health-in-2040-projected-patterns-of-illness-in-england (2023).

2. McGorry, P., Gunasiri, H., Mei, C., Rice, S. & Gao, C. X. The youth mental health crisis: analysis and solutions. Front. Psychiatry 15, 1517533 (2025).

3. Morris, S., Hill, S., Brugha, T. & McManus, S. Adult Psychiatric Morbidity Survey: Survey of Mental Health and Wellbeing, England, 2023/4. (2025) doi:10.13140/RG.2.2.24367.39840.

4. Department of Health and Social Care. Fit for the Future: 10 Year Health Plan for England. https://www.england.nhs.uk/long-term-plan/#:~:text=The%2010%20Year%20Health%20Plan,on%20The%20National%20Archives%20website (2025).

5. Muhl, C., Mulligan, K., Bayoumi, I., Ashcroft, R. & Godfrey, C. Establishing internationally accepted conceptual and operational definitions of social prescribing through expert consensus: a Delphi study. BMJ Open 13, e070184 (2023).

6. Oster, C., Skelton, C., Leibbrandt, R., Hines, S. & Bonevski, B. Models of social prescribing to address non-medical needs in adults: a scoping review. BMC Health Serv. Res. 23, (2023).

7. Bu, F., Hayes, D., Burton, A. & Fancourt, D. Equal, equitable or exacerbating inequalities: patterns and predictors of social prescribing referrals in 160 128 UK patients. The British Journal of Psychiatry 1–9 (2024) doi:10.1192/bjp.2024.141.

8. Drinkwater, C., Wildman, J. & Moffatt, S. Social prescribing. BMJ 364, 1–5 (2019).

9. Moscrop, A. Social prescribing is no remedy for health inequalities. BMJ 381, 715 (2023).

10. Calderón-Larrañaga, S., Milner, Y., Clinch, M., Greenhalgh, T. & Finer, S. Tensions and opportunities in social prescribing. Developing a framework to facilitate its implementation and evaluation in primary care: a realist review. BJGP Open 5, 1–13 (2021).

11. Melam, C., Dyson, J. & Thomson, K. The Role of Social Prescribing Interventions in Addressing Health Inequalities in the United Kingdom: A Narrative Review. Health Soc. Care Community 2025, (2025).

12. Cartwright L et al. Who Is and Isn’t Being Referred to Social Prescribing? https://socialprescribingacademy.org.uk/media/jaibqf4q/evidence-review-who-is-accessing-social-prescribing.pdf (2022).

13. Wilding, A., Sutton, M., Agboraw, E., Munford, L. & Wilson, P. Geographic inequalities in need and provision of social prescribing link workers a retrospective study in primary care. British Journal of General Practice 74, e784–e790 (2024).

14. Khan, K. et al. The feasibility of identifying health inequalities in social prescribing referrals and declines using primary care patient records. NIHR Open Research 3, 1 (2023).

15. Hayes, D. et al. Barriers and facilitators to social prescribing in child and youth mental health: perspectives from the frontline. Eur. Child Adolesc. Psychiatry 10.1007/s00787-023-02257-x (2023) doi:10.1007/s00787-023-02257-x.

16. Pescheny, J. V., Pappas, Y. & Randhawa, G. Facilitators and barriers of implementing and delivering social prescribing services: A systematic review. BMC Health Services Research vol. 18 Preprint at 10.1186/s12913-018-2893-4 (2018).

17. Bernard, K. et al. Experiences of Non-Pharmaceutical Primary Care Interventions for Common Mental Health Disorders in Socioeconomically Disadvantaged Groups: A Systematic Review of Qualitative Studies. International Journal of Environmental Research and Public Health vol. 20 Preprint at 10.3390/ijerph20075237 (2023).

18. Turk, A., Tierney, S., Hogan, B., Mahtani, K. R. & Pope, C. A meta-ethnography of the factors that shape link workers’ experiences of social prescribing. BMC Med. 22, (2024).

19. Jani, A., Liyanage, H., Okusi, C., Sherlock, J. & De Lusignan, S. Social Prescribing Observatory: A Learning Health System Approach for Using Data to Improve Practice. (2020).

20. NHS England. Neighbourhood health guidelines 2025/26. https://www.england.nhs.uk/long-read/neighbourhood-health-guidelines-2025-26/ (2025).

21. Gibson, K., Pollard, T. M. & Moffatt, S. Social prescribing and classed inequality: A journey of upward health mobility? Soc. Sci. Med. 280, (2021).

22. Bu, F. et al. National roll-out of social prescribing in England’s primary care system: a longitudinal observational study using Clinical Practice Research Datalink data. Lancet Public Health 10.1016/s2468-2667(25)00217-8 (2025) doi:10.1016/s2468-2667(25)00217-8.

23. NHS England. The NHS Long Term Plan. https://webarchive.nationalarchives.gov.uk/ukgwa/20230418155402/ https://www.longtermplan.nhs.uk/publication/nhs-long-term-plan/ (2019).

24. Mitchell, S. B. et al. The use of social prescribing and community-based wellbeing activities as a potential prevention and early intervention pathway to improve adolescent emotional and social development: a systematic mapping review. BMC Public Health 25, 3495 (2025).

25. Muhl, C. et al. Social Prescribing for Children and Youth: A Scoping Review. Health Soc. Care Community 2025, (2025).

26. Percy-Smith, B. ‘You think you know?…You have no idea’: Youth participation in health policy development. Health Educ. Res. 22, 879–894 (2007).

27. Kessler, R. C. et al. Lifetime Prevalence and Age-of-Onset Distributions of DSM-IV Disorders in the National Comorbidity Survey Replication. Arch. Gen. Psychiatry 62, 593–602 (2005).

28. YoungMinds & The Children’s Society. First Port of Call: The Role of GPs in Early Support for Young People’ s Mental Health. https://www.youngminds.org.uk/media/2csbkvlz/final-the-role-of-gps-in-early-support-for-young-peoples-mental-health.pdf (2021).

29. National Academy for Social Prescribing. Social Prescribing Link Worker Survey 2025 - Full Report. https://socialprescribingacademy.org.uk/resources/link-worker-survey-2025-understanding-what-matters-to-social-prescribing-link-workers/ (2025).

30. Elemental. https://www.theaccessgroup.com/en-gb/our-brands/elemental/.

31. Joy. https://www.thejoyapp.com/.

32. Social Rx Connect. https://www.socialrx.co.uk.

33. Bu, F., Hayes, D., Munford, L. & Fancourt, D. Assessing the impact of social prescribing on wellbeing outcomes: A national analysis of UK data. medRxiv 10.1101/2025.09.25.25336621 (2025) doi:10.1101/2025.09.25.25336621.

34. Wilding, A. et al. Have social prescribing referrals reduced appointment pressures on primary care? Evidence from Electronic Healthcare Records. British Journal of General Practice 75, bjgp25X741633 (2025).

35. Agboraw, E. et al. Inequality in the impact of the national social prescribing on population outcomes. British Journal of General Practice 75, bjgp25X741609 (2025).

36. Bone, J. K., Bu, F., Fancourt, D. & Hayes, D. Using electronic health records to evaluate a children and young people’s social prescribing service: Challenges and implications for research and practice. medRxiv 10.64898/2025.12.17.25342474 (2025) doi:10.64898/2025.12.17.25342474.

37. Benchimol, E. I. et al. The REporting of studies Conducted using Observational Routinely-collected health Data (RECORD) Statement. PLoS Med. 12, (2015).

38. Office for National Statistics. ONS Postcode Directory (May 2025) User Guide. https://geoportal.statistics.gov.uk/datasets/ca3396864fee4f5186046150f0e9092f/about (2025).

39. Scarpetti, G. et al. A comparison of social prescribing approaches across twelve high-income countries. Health Policy vol. 142 Preprint at 10.1016/j.healthpol.2024.104992 (2024).

40. Khan, H., Giurca, B. & International Social Prescribing Collaborative. Social Prescribing Around the World: A World Map of Global Developments in Social Prescribing Across Different Health System Contexts: 2024. https://socialprescribingacademy.org.uk/media/thtjrirn/social-prescribing-around-the-world-2024.pdf (2024).

41. Bradbury A et al. Social Prescribing in Child and Adolescent Mental Health Services: A Guide for CAMHS Practitioners. Learnings from Phase I of the UCL ‘Wellbeing While Waiting’ Study. https://sbbresearch.org/wp-content/uploads/2024/04/CAMHS-SP-guide-final.pdf (2024).

42. NHS England. Social Prescribing and Community-Based Support: Summary Guide. https://www.england.nhs.uk/wp-content/uploads/2020/06/social-prescribing-summary-guide-updated-june-20.pdf (2020).

43. NHS England. Primary Care Workforce Quarterly Update, 31 December 2025. https://digital.nhs.uk/data-and-information/publications/statistical/primary-care-workforce-quarterly-update/31-december-2025 (2026).

44. Bertotti, M. et al. Social prescribing for Not in Employment, Education or Training (NEET) young people: A realist evaluation of the C.O.P.E. project in Italy and Portugal. SSM - Mental Health 7, (2025).

45. Marques, M. J. et al. A realist synthesis of interventions for youth not in education, employment or training (NEET): building programme theories for effective support. International Journal of Adolescence and Youth vol. 30 Preprint at 10.1080/02673843.2025.2472022 (2025).

46. Pfeifer, J. H. & Peake, S. J. Self-development: Integrating cognitive, socioemotional, and neuroimaging perspectives. Dev. Cogn. Neurosci. 2, 55–69 (2012).

47. Sebastian, C. L., Burnett, S. & Blakemore, S.-J. Development of the self-concept during adolescence. Trends Cogn. Sci. 12, 441–446 (2008).

48. Howlett, N., Brown, K., Freethy, I., Mercer, S. & Özakıncı, G. We need better evidence for social prescribing: call for action for better systems for collaboration and building evidence. Perspect. Public Health 145, 135–137 (2025).

